# Detection and Recognition of Ultrasound Breast Nodules Based on Semi-supervised deep learning

**DOI:** 10.1101/2020.04.24.20078816

**Authors:** Yanhua Gao, Bo Liu, Yuan Zhu, Lin Chen, Miao Tan, Xiaozhou Xiao, Youmin Guo

## Abstract

**Background:** The successful application of deep learning in medical images requires a large amount of annotation data for supervised training. However, massive labeling of medical data is expensive and time consuming. This paper proposes a semi-supervised deep learning method for the detection and classification of benign and malignant breast nodules in ultrasound images, which include two phases.

**Methods:** The nodule position in the ultrasound image is firstly detected using the faster RCNN network. Second, the recognition network is used to identify the benign and malignant types of nodules. The method in this paper uses a semi-supervised learning strategy, using 800 labeled nodules and 4396 unlabeled nodules.

**Results:** Based on mean teacher training strategy, the proposed semi-supervised network has obtained excellent results, which is similar to currently used with supervised training networks. On the two test data sets, the AUC of semi-supervised learning and supervised learning were: 93.7% vs 94.2% and 92% vs 92.3%.

**Conclusions:** The paper proves that semi-supervised learning strategies have good application potential in medical images. Based on a special learning strategy, the result of semi-supervised learning is expected to achieve close or even achieve similar result of supervised deep learning, which only need a small number of labeled samples and a large number of unlabeled samples. It means deep learning analysis of breast lesion will be more feasible and more efficient.

## Background

Breast cancer is a common tumor in women worldwide. According to the 2019 American Cancer Report, breast cancer ranks first in new female cancer cases, accounting for about 30% of them in the United States [1]. The 2015 China Cancer Report pointed out that the incidence of breast cancer in Chinese women continues to increase, ranking first in women’s cancer [2]. Breast cancer can be manifested as palpable or non-palpable breast nodules, so the imaging diagnosis is of great significance for detecting and classifying indeterminate breast lesions as early as possible, and even improving breast cancer prognosis.

The imaging modalities includes breast magnetic resonance imaging (MRI), mammography, and breast ultrasound. Breast ultrasound plays a vital role in detecting breast nodules because of its convenience, no radiation, no contrast agent is needed and cost-effectiveness. Moreover, ultrasound can be more sensitive to detect occult breast tumors in dense-breasted women compared with mammography [3]. Although ultrasound breast examination has good sensitivity and specificity, it may be affected by certain factors such as doctor’s personal experience. Moreover, it is difficult to classify the breast nodules of some atypical malignant signs. Usually, the ultrasound examination is done with by single sonographer in China. This diagnostic process is subjective, monotonous and tired, prone to errors. Artificial intelligence has been proposed to assist sonographers. There is room for making the diagnosis of breast cancer more reliable and efficient.

In recent years, deep learning has been widely used in medical image analysis, such as CT [4], MRI [5] and X-ray [6]. Deep learning has also made a lot of progress in ultrasound images, involving classification [7-10], image quality assessment [11,12], standard plane detection [13], image registration[14], automatic image segmentation[15, 16], removal of sensitive information [17], image alignment[18],disease assessment[19]. The end-to-end training in deep learning, which automatically extracts representations and features in the image, may lead to expert-level identification methods of diseases. Based on the pre-trained GoogleNet model, fine-tuned train to identify the benign and malignant thyroid nodules, achieves 90% of accuracy [20]. Deep learning to assess patients with myocarditis, distinguishing normal, infection and myositis, is more accurate than traditional methods [21]. Other studies include the recognition of thyroid follicular adenomas and cancers [22], detection of aortic stenosis [23], location of thyroid nodules [24]. Deep learning of animal ultrasound even detects degenerative liver disease in dogs [26].

Deep learning analysis of breast images has yielded many valuable results. Convolutional neural networks were used to detect the location of breast lesions [27], and classify the benign and malignant breast nodules in CT images, breast lesions in ultrasound images and so on. Based on deep learning, automatic detection and diagnosis of breast cancer are possible, and find four different pathological classifications of breast lesions [28-31].

Deep learning has made great progress in diagnosing breast nodules, but almost all deep learning models are supervised [32]. Supervised deep learning methods are mostly based on neural networks, using backward diffusion algorithms to train weights. The performance of deep neural networks is generally related to capacity, and the deeper and wider network architecture with large number of parameters is often chosen to ensure performance. In order to make the network converge, a large amount of manpower is required to manually label the location and pathological result of the nodule [33]. Although the hospital has a large amount of breast nodule images, the labeled data is very limited.

Ideally, the unsupervised deep learning method can automatically learn the representation in the image and realize various tasks such as image segmentation, recognition, detection, video tracking [33]. However, the research of such methods is difficult. Compared with the supervised method, unsupervised method has poor performance, difficult to use in practical application. The semi-supervised learning methods requiring only a small number of labeled samples, together with a large number of unlabeled samples, are expected to achieve near or even better than results of supervised methods, and has great potential for development, especially for medical application.

This paper combines the Faster RCNN network [34] and mean teacher semi-supervised learning method [35]. It includes a two-stage framework to realize semi-supervised detection and recognition of breast nodules in ultrasound images. The paper has the following highlights:

1. As far as we know, this paper is the first semi-supervised method for identification of benign and malignant breast nodules. With only a few hundred labeled breast nodules, the proposed method can obtain similar accuracy of supervised learning.
2. The paper demonstrates the potential of semi-supervised deep learning methods in medical images, which is expected to greatly reduce the pressure of data annotation and speed up application in medical image analysis.

## Materials and Methods

### Dataset

The dataset comes from two independent hospitals, The Third Affiliated Hospital of Xi’an Jiaotong University and The First Affiliated Hospital of Xi’an Jiaotong University. The study was approved by relevant Institutional Review Boards, and a general research authorization was obtained allowing for retrospective reviews. Informed consent was waived by IRB. From the hospital information system, the technicians retrieved the patients who entered the hospital for ultrasound breast examination and had pathological diagnosis in 2010-2019. In order to ensure the representativeness of the data, the number of patients in each week does not exceed 50 cases, and 9012 nodules images from 9012 patients were obtained. The data retrieval task was performed by the information system administrator, and the ultrasound doctors and pathologists were not involved in data collection. The technician did not check the image quality, and did not participate in the follow-up study. The data of the two hospitals were taken as dataset A and dataset B respectively.

The data set A was from The Third Affiliated Hospital of Xi’an Jiaotong University, and data set B was from The First Affiliated Hospital of Xi’an Jiaotong University.

### Dataset review

The breast nodule images were obtained from data sets A and B, and two ultrasound doctors with 10 years’ clinical experience reviewed them to check image quality. The labeling tool is used to label the position of breast nodule by a bounding box. The rule is to include all nodule tissue and the background tissue of approximately 10 pixels. According to the pathological diagnosis, the bounding box is given with benign or malignant type at the same time. The two doctors were no longer involved in the follow-up study.

After image review, some images with poor image quality were removed, and the two data sets retained 8966 nodules. Among them, there were 6746 nodules in dataset A, 2809 malignant (breast cancer) and 3937 benign (non-breast cancer). Dataset B has 2,220 nodules, which was used as an independent test set, including 1291 malignant nodules and 929 benign nodules.

### Train, validation and test datasets preparation

Dataset A is used to train and test neural networks. In order to perform semi-supervised learning, dataset A was randomly divided into training set (about 15%), extended training set (65%), and testing set (20%). The training set consists of 500 malignant and 500 benign nodules, which is divided into basic training set (80%) and validation set (20%). The basic training set is less than one-fifth of the extended training set. The basic training set uses fully annotated information, including the location of bounding box and the pathological results, malignant and benign, respectively.

Although the extended training set has been labeled with the bounding box and breast types, the annotated information is not used in the following semi-supervised learning. The validation set used 100 malignant and benign nodules for hyperparametric adjustments in training. The test set accounts for 20% of dataset A to test the trained model. Dataset B serves as an independent test set to verify the generalization capabilities of the method.

### The framework of the network

Considering that the data in basic training set is very small, this paper divides the detection and recognition into two phases. The first stage only achieves the positioning of breast nodules, and does not distinguish between benign and malignant categories, in order to speed up the training of the network. The second stage identifies malignant and benign of breast nodules in the bounding box.

Two networks are performed to complete the tasks of these two stages, i.e. detection network, and recognition network. The detection network is to detect the position of bounding box including breast nodules in ultrasound image. The faster RCNN network is used for detection [34], while the feature extraction part is the VGG 16 network [36], and the region proposal network is used to output the possible bounding box. The faster RCNN outputs two categories, namely the foreground (breast nodule), the background (curves, text, color bars, etc.), as shown in Figure 1.

**Figure 1.**
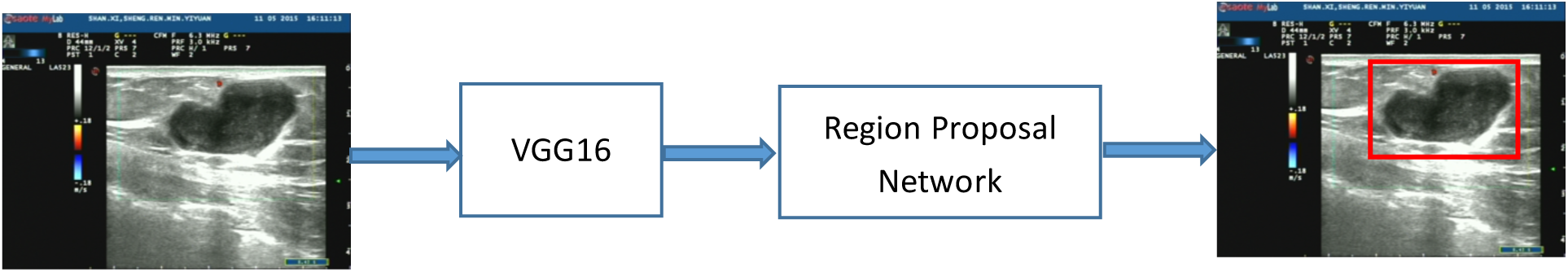
Faster R-CNN identifies the position of bounding box surrounding the nodules. An original ultrasound image is input into the VGG 16 to acquire image features. Region proposal network presents some possible bounding box.

The bounding box of breast nodules is obtained by faster RCNN, and the picture in the box is sent into recognition network. The recognition network uses a semi-supervised learning strategy called mean teacher, including a student network and a teacher network, as shown in Figure 2. The student network learns both labeled images (basic data sets) and unlabeled images (labeled by teacher network), and define recognition errors as the joint cost function [35]. The student network and the teacher network have the same network architecture, including eight convolution layers and some pooling layers and full connection layers. The student network includes three modules, and each module includes three convolution layers, a maximum pooling and dropout layer. Finally, two convolution layer, one average pooling and fully concoction layer are performed for output.

**Figure 2.**
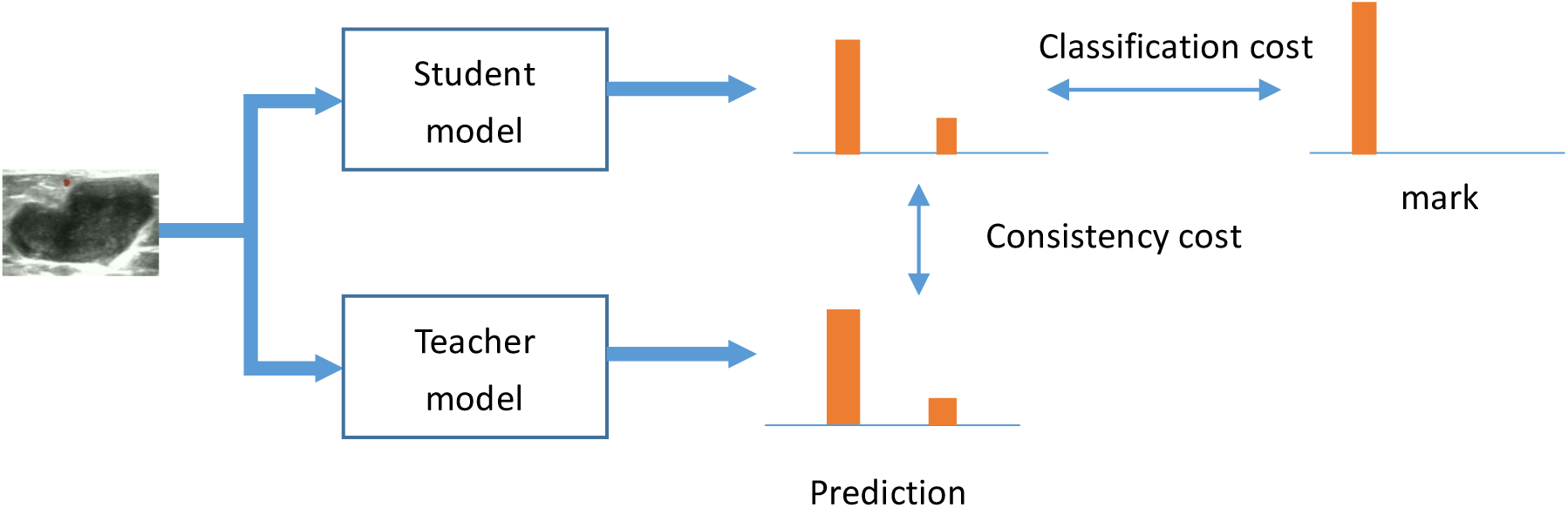
recognition network, the semi-supervised learning of mean teacher. The student network uses softmax to get the one-hot label of the output. If the input sample has a label, calculate the classification cost. If there is no label, the consistency between the student and teacher is considered as the cost function. After the weights of the student model are updated, the weights of the teacher model are updated by the exponential moving average of the student weight. The output of both models can be used as a prediction, but the results predicted by the teacher are more likely to be correct.

### Network training

#### (1) Faster RCNN re-training in ultrasound images

The 800 ultrasound images and bounding box of breast nodules in basic training set were entered to learn the position of the nodules. In order to speed up the training, the faster RCNN network is initialized using a pre-trained model on the ImageNet dataset, and fine-tuning training is performed in the next step. Firstly, shallow fine-tuning faster RCNN, that is, the feature extraction layers by the VGG16 network are frozen, while the region proposal layers at which the bounding box is generated are trained. After 5000 iterations, the VGG16 is allowed to retrain weights for deep fine-tuning. The hyperparameters have been proven to achieve satisfactory positioning accuracy by validation sets. The hyperparameters are: initial learning rate of 0.001, decreases exponentially, momentum of 0.9. The batch size is 128, optimizer is momentum optimizer, and total number of iterations is 70,000.

#### (2) Training the recognition network

The images in basic training set and extended training set are input into fast RCNN to obtain bounding boxes, and the nodule images inside the boxes are extracted. The labels of nodule from basic training set are known, but labels from extended training set are unknown and provided by teacher network. The semi-supervised mean teacher method is used to train the teacher and student network. The optimized hyperparameter is: batch size is set to 128, in which 64 images are randomly taken from basic training set, but another 64 are randomly from extended training set. The kernels of the convolutional layer are initialized by Gaussian with a mean of 0, a standard deviation of 0.05, and then L2 norm. The cost function consists of two parts. The first is the classification error, calculated by the cross-entropy of the true labels and the predicted labels of basic training set. The second is the consistency error, which is used for the unlabeled breast nodules to measure the mean square error between the predicted labels of student and teacher.

The student model is trained with the teacher model, where the teacher model is updated in an exponential moving average manner. The data augmentation such as image flipping, random change in brightness is used. Other hyperparameters are: learning rate is set to 0.003, input noise is 0.15, and dropout ratio is 0.5. The optimizer is Adam, and the number of iterations is 40,000. The model with best accuracy on validation set is retained, at the same time, the early stopping strategy is used, when there are 5 consecutive epochs in the validation set that accuracy no longer improves, the training will be stopped.

## Results

After finishing the training of the two networks, the results are validated on dataset A and dataset B. The results include detection accuracy and sensitivity and specificity of classification, which are produced by two networks. The classification is based on bounding box provided by the detection network to obtain benign or malignant type.

### (1) Detecting position of breast nodules

This paper uses the average similarity criterion(ASC) to indicate the detection accuracy, which is defined as (*R*_1_ ∩ *R*_2_)/(*R*_1_ ∪ *R*_2_), where *R*_1_ is the bounding box of one nodule given by ultrasound doctors, but *R*_2_ is predicted bounding box of the nodule. The mean and standard deviation of ASC results are shown in Table 2.

**Table 1.**
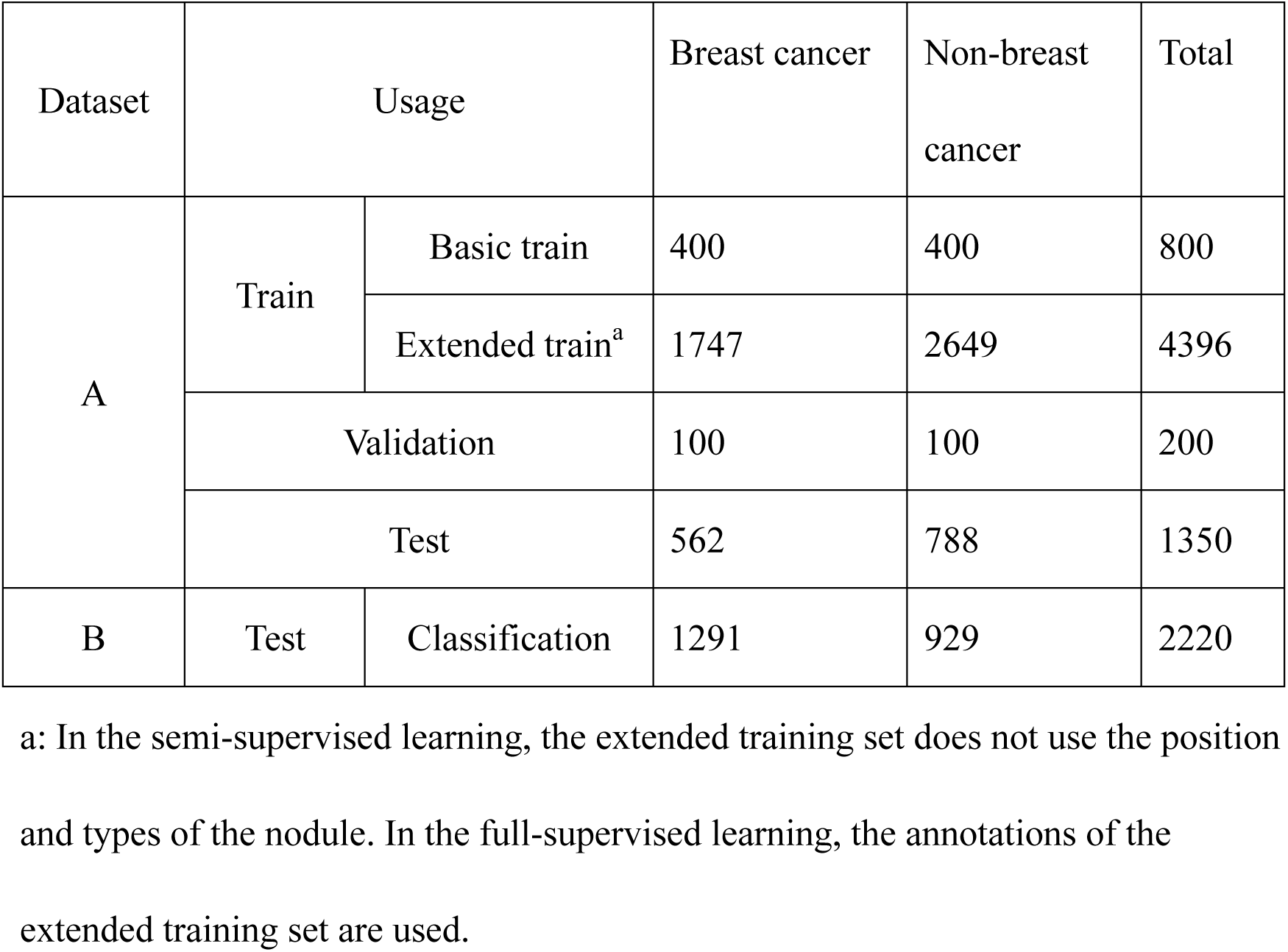
Datasets of breast nodules

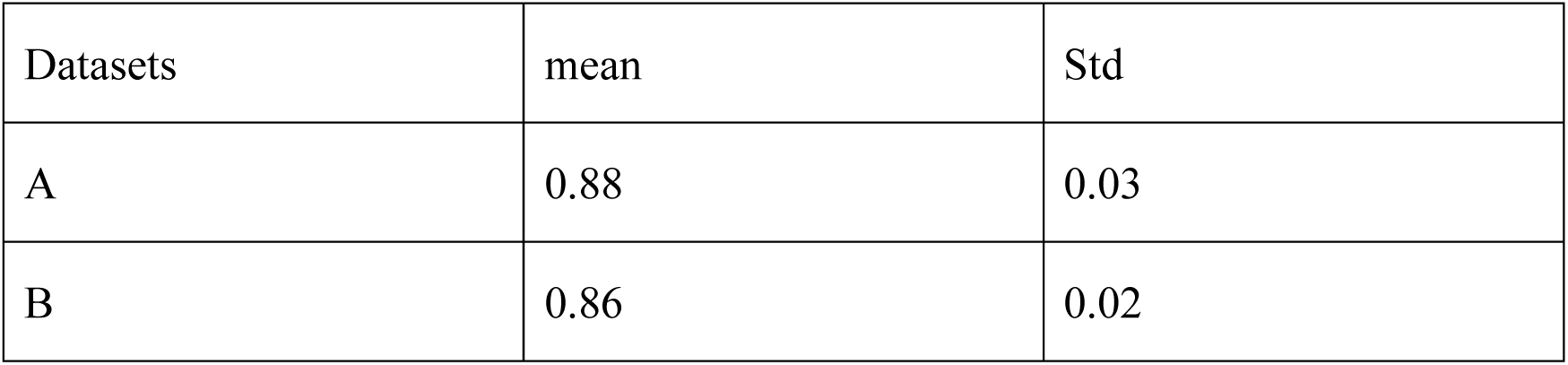

| Datasets | mean | Std |
| --- | --- | --- |
| A | 0.88 | 0.03 |
| B | 0.86 | 0.02 |

As can be seen from Table 2, the mean value of ASC is above 0.85, indicating that the detection network is very consistent with the doctor, and can be used for detecting the position of the nodules.

Figure 3 shows nodule localization on some images, where the yellow bounding box represents the position given by the doctor and red is given by the detection network. As can be seen from the degree of overlap of the two bounding boxes, the Faster R-CNN network can achieve a sufficiently accurate detection of the breast nodules in the images.

**Figure 3.**
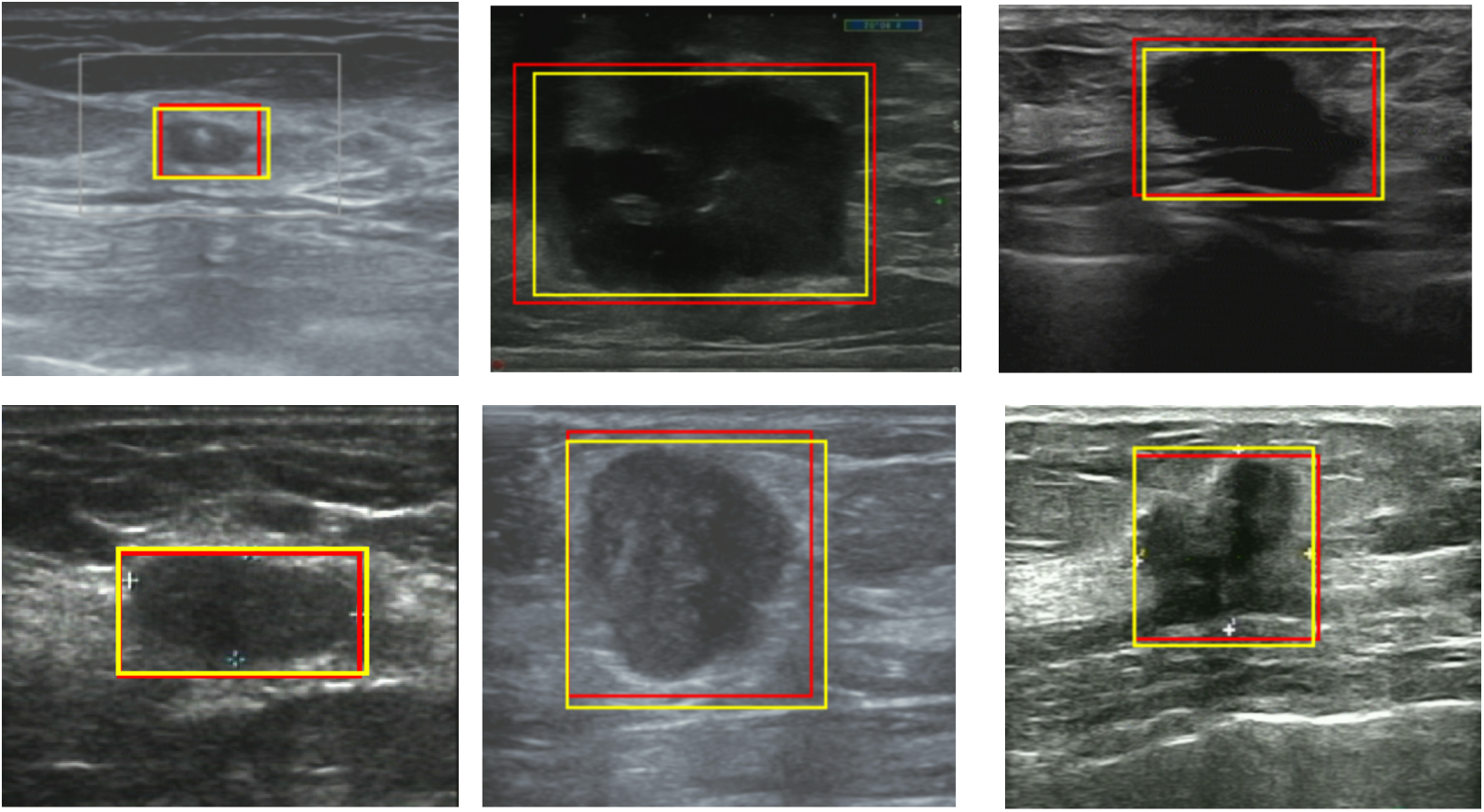
Localization results of breast nodules. In order to achieve a better display, the rest of the images away from the nodules were removed.

### (2) The classification of benign or malignant type

The sensitivity, specificity, accuracy, and AUC (receiver operating characteristic curve) value are used to validate the classification. Sensitivity is defined as the number of correctly classified malignant nodules divided by the number of all malignant nodules, and specificity is defined as the number of correctly classified benign nodules divided by the number of all malignant benign nodules. The accuracy is defined as the number of correct classified nodule divided by the total number of all nodules; the AUC value is defined as the area under the ROC curve.

For comparison, we train two networks of supervisors learning respectively, which use basic training set and all training set including basic training set and extended training set. The network structure with supervised learning is the same as the student network of semi-supervised learning. The validation set is used for selecting hyperparameters to obtain the optimized results. The classification results of dataset A and B are shown in Table 3 and 4.

**Table 3.** Test results of data set A

| Methcod | Train datasets |  | Sensitivit<br>y | Specificit<br>y | Accuracy | AUC |
| --- | --- | --- | --- | --- | --- | --- |
|  | with<br>marks | Without<br>marks |  |  |  |  |
| Semi-sup<br>ervisor | A-basic<br>train | A-extra<br>train | 87.9% | 89.7% | 88.8% | 93.7% |
| Superviso<br>r<sup>a</sup> | A-basic<br>train | / | 72.2% | 75.1% | 73.8% | 83.3% |
| supervisor<br><sup>b</sup> | A-basic<br>train+A-e<br>xtra train | / | 89.1% | 90.2% | 89.6% | 94.2% |

The supervisor<sup>a</sup> is the classification results of detection network trained by supervised methods using 800 marked nodules and 100 validation sets. The supervisor<sup>b</sup> used 5396 nodules from the two training sets for supervised learning, where benign and malignant labels in extended training set are used. The semi-supervisor is the classification results of this presented method, where the 800 labeled nodules and 4396 unlabeled nodules are used for mean teacher learning.

The accuracy of Supervisor^a^ is about 73.8%, which is lower than the other two methods. It shows that the basic training set is too small to extract enough representative information and the network is easy to over-fitting and performs poorly on the test set. The supervisor^b^ network is trained by all training data, whose classification accuracy is the best, and similar to that of the Semi-supervisor. The result indicates the semi-supervised method using only one-fifth of the labeled image along with other unlabeled images, may obtain the similar accuracy to that of supervised learning using all labeled nodule images. This result illustrates possible potential medical application of semi-supervised learning.

Table 4 shows the results of the independent test set B. Compared with Table 3, the accuracy is slightly reduced, which shows that the method of semi-supervised learning also has good generalization ability. Similarly, the results of semi-supervision are closer to those of supervisor learning.

**Table 4:** Results of Independent Test Set B

| Method | Train datasets |  | Sensitivit<br>y | Specificit<br>y | Accuracy | AUC |
| --- | --- | --- | --- | --- | --- | --- |
|  | with<br>marks | Without<br>marks |  |  |  |  |
| Semi-sup<br>ervisor | A-basic<br>train | A-extra<br>train | 85.3% | 86.8% | 85.9% | 92.0% |
| Superviso | A-basic | / | 73.2% | 70.1% | 71.9% | 81.8% |
| $r^a$ | train | | | | | |
| supervisor<br>$b$ | A-basic<br>train+A-e<br>xtra train | / | 87.3% | 86.5% | 86.9% | 92.3% |

### (3) Compare with the results of an ultrasound doctor

In order to better illustrate the results of the semi-supervised method, the 5 experienced ultrasound doctors identified benign and malignant types of independent test set B. The clinical experiences of the 5 ultrasound doctors are shown in Table 5.

**Table 5.** clinical experiences of 5 ultrasound doctors

| ultrasound doctors | clinical experiences |
| --- | --- |
| A | 3 years |
| B | 4 years |
| C | 7 years |
| D | 10 years |
| E | 15 years |

**Table 6.** classification results of an ultrasound doctor and artificial intelligence

|  | AI | Ultrasound doctors |  |  |  |  |  |
| --- | --- | --- | --- | --- | --- | --- | --- |
|  |  | A | B | C | D | E | average |
| Sensitivity | 85.3% | 76.5% | 73.2% | 79.8% | 85.3% | 82.5% | 79.5% |
| specificity | 86.8% | 72.4% | 79.4% | 82.4% | 89.5% | 80.5% | 80.8% |
| accuracy | 85.9% | 74.7% | 75.7% | 80.9% | 87% | 81.6% | 79.9% |
| AUC | 92.0% | 83.9% | 86.6% | 89.9% | 93.6% | 90.3% | 88.9% |

The AI(artificial intelligence) column is the sensitivity, specificity, accuracy and AUC of presented semi-supervisor method. The final column is the average result of 5 doctors. Compared with the average result, AI is more accurate than humans, where the accuracy and AUC of AI and doctors are 85.9% vs. 79.9% and 92.0% vs. 88.9% respectively.

With the acceleration of the Nvidia Telsa V100 GPU, the identification of dataset B is completed in just a few seconds, much faster than the speed of the ultrasound doctor. In order for the ultrasound doctors to estimate the benign and malignant nodules as accurately as possible, there is no timing in this experiment. The results are shown in Table 6.

## Discussion

In recent years, deep learning has been extensively studied in medical images and has achieved impressive accuracy. Deep learning is based on a multi-layer neural network. Through multiple nonlinear transformations, the relationship between input data and output results is obtained. However, for deep learning to achieve good results, there must be a large amount of labeled data. The networks trained by a small number of labeled data are prone to overfitting and poor generalization on independent test sets.

Labeled medical data is often very expensive. The amount of clinical data is always very large, but it lacks manual marking. The task of labeling data requires many medical experts and takes a lot of time. The use of unlabeled data to train deep neural networks with unsupervised methods is still under study, and the accuracy is far from practical applications. Therefore, it is promising to use a small amount of labeled data along with a large number of unlabeled data in medical data processing, which is semi-supervisor learning.

The detection and classification of breast nodules is a typical ultrasound task. Previous studies have adopted supervised learning strategies, using thousands or even more nodules as training sets, which have achieved good results in some datasets. Such a data set not only needs to label benign and malignant, but also needs to know pathological confirmation of the nodule, which takes a lot of time to obtain. More importantly, in order to improve nodule recognition from multi-center data, more labeled data is required.

This paper presented a semi-supervised learning model for the detection and recognition of ultrasound breast nodules. The model is divided into two stages, first the faster RCNN network is used to provide possible position of breast nodules. Because of the relatively small number of labeled nodules in training set, the faster R-CNN network only recognizes two categories, nodules and backgrounds. The results have shown the faster RCNN network trained on ImageNet through transfer learning with a small number of hundreds of labeled nodules, Identifies the location of the nodule, and well identifies the interference information such as text, color bars in ultrasound images.

The mean teacher networks consist of two models, student network and teacher network. For labeled samples, the cost is calculated by predicted label and real label provided by doctors, and for the unlabeled samples, the consistency between teacher and student is used for cost function. The teacher network is equivalent to a smooth version of student network and is less prone to overfitting. It is possible that the accuracy of semi-supervisor learning on a small amount of labeled data along with a large number of unlabeled data sets, is close to that of supervised learning using all labeled data.

The test set and training set in dataset A are from the same hospital, and the semi-supervised results are satisfactory, which indicate that the presented method can achieve good recognition result. Dataset B is an independent test set from another hospital, and the method has good generalization ability too. The semi-supervised learning using about one-fifth of the labeled data can obtain similar results with supervised learning strategies, indicating the potential application of semi-supervised methods in medical data.

AI and human doctors also made a comparison. There were five doctors of different expectances who participated in the competition, and they did not participate in the model training. The results of AI and humans showed that the method ranked second among the six testers (including AI), higher than the doctor’s average (accuracy 85.9%: 79.9%). This result proves that the deep learning network based on semi-supervised learning can also achieve the same accuracy or even better than that of ultrasound doctors.

The highlight of this paper is the use of semi-supervised learning strategies to train the detection and classification of ultrasound breast nodules, which proves that the method can achieve the same accuracy as supervised learning and can reach the level of ultrasound doctors. The semi-supervised learning strategy reduces the number of labeled data, and it is easier to be applied in medical applications.

This paper confirms that based on hundreds of labeled nodules, along with thousands of unlabeled nodules, the results of semi-supervisor learning are close to that supervised learning, and achieve good results on independent test sets. This method is expected to be applied in other medical images, such as pathological images, magnetic resonance images, X-ray images and so on. The recognition is a basic problem, and accurate recognition can help solve other problems, such as prediction of treatment outcomes.

## Conclusion

This paper proposed a detection and classification method of breast nodules based on semi-supervised learning, which is trained on a small amount of labeled data and is close to the accuracy of supervised learning. The work of this paper is not only helpful to build the recognition model of breast nodules, but also be extended to other medical images and data. The reveals the application prospects of semi-supervised learning, which helps to solve the costly and time-consuming problems of current medical data annotation.

## Data Availability

The data that support the findings of this study are available from The First Affiliated Hospital of Xi’an Jiaotong University and The Third Affiliated Hospital of Xi’an Jiaotong University but restrictions apply to the availability of these data, which were used under license for the current study, and so are not publicly available.

## Abbreviations

RCNN: Regional Convolutional Neural Network features

## Ethics approval and consent to participate

Obtained on the study is included in the “Methods” section.

## Consent for publication

Not applicable.

## Funding

Not applicable.

## Competing interests

The authors declare that they have no competing interests.

## Contributions

All authors read and approved the final manuscript. YM. Guo was responsible for the overall study supervision and reviewing manuscript. YH. Gao was responsible for the study design and was a major contributor in writing the manuscript. YH. Gao, B. Liu, Y. ZH, L. CH, and M.T were responsible for the data collection (imaging acquisition, image analysis, image interpretation, clinical and pathologic outcomes). X.Z Xiao were responsible for the statistical analysis and interpretation of the data.

## Acknowledgements

Not applicable.

## Notes

### Competing Interest Statement

The authors have declared no competing interest.

